# The Rural Adversity and Determinants Index: A Novel Assessment Method of At-Risk Rural Counties

**DOI:** 10.1101/2025.10.10.25337725

**Authors:** Eashwar S.C. Krishna, Apurv Manjrekar, Nicolas Araujo, Quang Dai La

**Affiliations:** Department of Sociology, University of Connecticut, Storrs, United States; Department of Computer Science, University of Connecticut, Storrs, United States; Department of Biochemistry and Molecular Biology, University of Massachusetts Amherst, Amherst Center, United States; Department of Biology, Texas A&M University, College Station, United States

## Abstract

**Introduction:** Significant rural health disparities are driven by social determinants of health (SDOH), yet existing tools like the CDC’s Social Vulnerability Index (SVI) and USDA’s Rural-Urban Continuum Codes (RUCC) inadequately capture rural-specific adversity due to national-level aggregation and geographic oversimplification. This study develops and validates the Rural Adversity and Determinants Index (RADI), a novel composite SDOH index tailored for U.S. rural counties. We hypothesized RADI would outperform SVI and RUCC in predicting adverse health outcomes and uniquely explain rural health disparities.

**Methods:** Using 1,210 non-metropolitan U.S. counties (RUCC >3), RADI was constructed via Principal Component Analysis from 18 SDOH metrics spanning income, education, infrastructure, and healthcare access. Validation against five health outcomes (premature mortality, poor/fair health, infant mortality, heart disease mortality, preventable hospital stays) included bivariate correlations, single-predictor linear regressions (comparing RADI, SVI, and RUCC via R^2^), and multivariable regressions testing RADI’s unique contribution after controlling for SVI/RUCC.

**Results:** RADI significantly outperformed SVI and RUCC across all outcomes. For premature mortality, RADI explained 55.6% of variance versus SVI (37.3%) and RUCC (2.4%); for poor/fair health, RADI R^2^ reached 77.2% versus SVI (50.1%). RADI correlated strongly with SVI (r = 0.708) but captured distinct adversity dimensions: RADI scores increased with geographic isolation (β = 0.0278, p<0.001 for RUCC), while SVI unexpectedly decreased (*β* = –0.4279, p=0.024) at high rurality. In multivariable models, RADI remained a highly significant predictor after controlling for SVI/RUCC (e.g., for premature mortality: *β* = 95.29, p<0.001), whereas RUCC lost significance for most outcomes.

**Conclusions:** RADI is a rigorously validated, rural-specific SDOH index that more accurately quantifies adversity than national tools. By exposing modifiable drivers like infrastructure deficits and healthcare access gaps, RADI enables precise targeting of policies and resources to reduce rural health inequities. Future work should integrate RADI into funding frameworks and expand its use in longitudinal and intervention studies

## 1.0 Introduction

### 1.1 The Landscape of Rural Health Disparities

It is well-documented that significant health disparities persist between urban and rural populations within the United States [1]. Frequently, rural communities are confronted with a set of distinct and significant challenges. Among these challenges is often limited access to a range of healthcare services, which includes both primary and specialty care [2]. Residents of rural areas also tend to experience higher rates of chronic disease and premature mortality when compared to their urban counterparts [3]. These health challenges are compounded by underlying socioeconomic factors, such as economic instability and various infrastructure deficits [4–5]. For instance, such infrastructure issues can include limited access to broadband internet and a lack of reliable transportation, which in turn can further isolate residents from essential services [4–5].

To understand these complex issues, it is necessary to use a framework that looks beyond the scope of clinical care. The Social Determinants of Health (SDOH) framework is ideal for this purpose. This concept illuminates how the conditions in the environments where people are born, live, learn, work, and age can affect a wide range of health outcomes and risks [6]. Key factors like income, level of education, and housing quality serve as powerful predictors of overall health status [7]. For rural areas specifically, the SDOH framework offers a critical lens that can be used to analyze and address the root causes of long-standing health inequities [8]. Therefore, maintaining a focus on SDOH is essential for the development of effective policies and interventions that are tailored to the unique circumstances of rural communities throughout the nation [8].

### 1.2 The Need for Nuanced Measurement: Limitations of Existing Indices

To identify and provide assistance to vulnerable populations, researchers and policymakers often rely on national indices [9]. These tools, however, have significant limitations when applied to rural contexts due to their broad, national-level focus [9]. One widely used measure is the Social Vulnerability Index (SVI) from the Centers for Disease Control and Prevention (CDC) [10]. While the SVI is a valuable tool for assessment at a national level, it can prove insufficient for the purpose of understanding rural adversity [11]. Its data aggregation methods often mask important “within-rural” variations. The population of counties designated as “rural” is not a monolith; instead, it represents a wide spectrum of socioeconomic conditions. The SVI may fail to capture the specific drivers of adversity that are unique to the most remote or distressed of these areas, which could potentially lead to an inefficient allocation of resources [12]. Furthermore, evidence exists to suggest that the SVI may “break down” in rural contexts, displaying a bell-curve pattern such that extremely rural areas are mistakenly shown to have lower vulnerability than suburban/rural mixed areas [13]

Limitations are also present in purely geographic classifications. The Rural-Urban Continuum Codes (RUCC) from the U.S. Department of Agriculture (USDA) are useful for categorizing counties based on their population size and proximity to metropolitan areas. This system is effective in separating urban and rural counties into distinct groups. However, the RUCC system itself does not inherently measure socioeconomic conditions, access to resources, or the lived experience of adversity. A county’s geographic classification, by itself, provides little information about the health, economic stability, or critical infrastructure access of its residents. It is possible for two counties with the exact same RUCC designation to have vastly different levels of social and economic well-being. This situation creates a critical gap in both the academic literature and in practical application. A clear need exists for a new measurement tool – an index that moves beyond simple geographic classification in order to quantify the degree of adversity across the full spectrum of rurality.

### 1.3 Project Aims and a New Hypothesis: The Rural Adversity and Determinants Index (RADI)

The present project seeks to address the aforementioned gap through the development and validation of a novel measurement tool. Specifically, the primary objective of this study is the development and validation of a new, composite Social Determinants of Health (SDOH) index that is tailored to the unique context of U.S. rural counties. We have given this tool the name Rural Adversity and Determinants Index (RADI). To create a more granular measure of vulnerability within rural America, the RADI is constructed from eighteen distinct metrics that span multiple domains, including income, education, health access, and infrastructure.

We have established several secondary objectives for the validation of the RADI. First, an assessment of the RADI’s predictive validity against key population health outcomes, such as premature mortality and the prevalence of poor or fair health, will be conducted. Second, we will perform a direct comparison of the predictive power of the RADI with that of the CDC’s SVI and the USDA’s RUCC classifications. Third, our study will determine if the RADI provides unique explanatory power for health outcomes that remains even after accounting for the influence of the SVI and RUCC.

This investigation is guided by three central hypotheses: **H1:** The Rural Adversity and Determinants Index (RADI) score will be significantly and positively correlated with adverse population health outcomes in U.S. rural counties. **H2:** The RADI will explain a greater proportion of the variance in these adverse health outcomes than either the SVI or the RUCC system can explain alone. **H3:** The RADI will remain a statistically significant predictor of adverse health outcomes, even when controlling for the effects of SVI and RUCC in multivariate models.

## 2.0 Methodology

### 2.1 Study Design and Scope

This study employed a cross-sectional ecological design to develop and validate a novel composite index of rural adversity. The unit of analysis for all data collection and subsequent statistical modeling was the U.S. county. This level of aggregation is appropriate for examining the broad social, economic, and environmental factors that shape population health outcomes and allows for the integration of data from various national surveillance systems.

The study sample was intentionally focused on non-metropolitan counties to ensure the resulting index was specifically tailored to rural contexts. The sample comprises 1,210 U.S. counties that are classified as rural, defined as having a 2023 U.S. Department of Agriculture (USDA) Rural-Urban Continuum Code (RUCC) greater than 3. The RUCC scale ranges from 1 (most urban) to 9 (most rural). The cutoff of RUCC > 3 was selected to exclude all metropolitan counties (codes 1-3), thereby focusing the analysis exclusively on counties ranging from “Urban population of 20,000 or more, adjacent to a metro area” (RUCC 4) to “Completely rural or less than 2,500 urban population, not adjacent to a metro area” (RUCC 9). This specific definition ensures the index captures adversity across the entire rural continuum, from large rural towns to the most isolated non-metropolitan regions.

### 2.2 Data Sources and Variable Selection

To construct a comprehensive index, data were drawn from multiple authoritative national sources, each chosen for its relevance to specific social determinants of health. These sources included the U.S. Census Bureau’s American Community Survey (ACS) for foundational data on income, poverty, employment, and housing; the HHS/HRSA Area Health Resource Files (AHRF) for critical data on healthcare professionals and resources; the University of Wisconsin Population Health Institute’s County Health Rankings (CHR) for validation outcomes and metrics like food insecurity; the Environmental Protection Agency (EPA) for data on drinking water violations; the U.S. Department of Agriculture (USDA) for geographic classification; and the Federal Emergency Management Agency (FEMA) for environmental risk data [14–18].

From these sources, 18 distinct metrics were selected to create a multidimensional view of rural adversity:

- Household median income
- % in poverty 18-64
- % unemployed
- % lacking complete plumbing facilities
- % under housing cost burden
- % without internet
- % with no vehicle available
- % with long commutes
- Drinking Water Violations (binary)
- % of population aged 65+
- % non-White population
- % less than high school graduate
- % without some college or associate’s degree
- % Uninsured population
- Ratio of population to primary care physicians
- Mental Health Provider rate (inverted)
- % experiencing food insecurity
- Natural Disaster Risk score

### 2.3 Index Construction: The RADI Algorithm

The construction of the RADI followed a systematic process executed primarily in Python; this process began with data preparation, where county-level data for the 18 selected metrics were aggregated and merged into a single dataset using the pandas library, aligning counties by their FIPS codes. To handle the small fraction of missing data (0.53%), we used an iterative imputer, an advanced method that preserves multivariate relationships by modeling each feature as a function of others. Following imputation, all 18 metrics were Z-normalized to have a mean of 0 and a standard deviation of 1. This normalization is critical for PCA, as it prevents variables with larger scales from disproportionately influencing the analysis. Variables were also re-oriented so that higher values consistently indicated greater adversity; median income and mental health provider rate data were inverted by multiplying by negative one. The core of the index construction was Principal Component Analysis (PCA), a dimensionality reduction technique used to combine the 18 correlated variables into a single composite score. We implemented PCA to extract the first principal component (PC1); this raw PC1 score was transformed into the final RADI score by scaling it to an intuitive 0-100 range, where 0 signifies the lowest and 100 signifies the highest level of adversity in the rural sample.

Principal Component Analysis (PCA) was deliberately chosen as the construction method for the RADI over other approaches like expert-defined or equal weighting. The primary advantage of PCA is its data-driven objectivity; it avoids the subjectivity inherent in predetermined weighting schemes by algorithmically calculating variable weights based on the shared patterns within the data itself. The goal was to create a single, parsimonious index that captures the most significant underlying dimension of adversity shared across the 18 metrics. By using the first principal component (PC1) – which, by definition, explains the maximum possible variance – we ensure the resulting RADI score is the most faithful composite summary of the data’s primary dimension. While other methods like Factor Analysis exist, PCA was selected for its directness as a dimensionality reduction technique ideally suited for creating a descriptive and summary index without imposing a stricter causal modeling framework.

### 2.4 Validation Strategy and Statistical Analysis

A multi-stage validation strategy was designed to rigorously assess the RADI’s utility and performance. The RADI’s predictive validity was tested against five key population health outcomes from the County Health Rankings dataset: Poor / Fair Health (percentage of adults reporting fair or poor health), Premature Mortality (years of potential life lost before age 75 per 100,000 population), Infant Mortality (deaths per 1,000 live births), Heart Disease Mortality (deaths per 100,000 population), and Preventable Hospital Stays (hospitalization rate for ambulatory-care sensitive conditions per 100,000 Medicare enrollees).

We began with a bivariate analysis, calculating Pearson correlation coefficients to measure the linear relationship between the RADI score and each health outcome. Next, to assess comparative predictive power, a series of single-predictor linear regression models was run to quantify the proportion of variance (R^2^) in each health outcome explained independently by RADI, SVI, and RUCC. We then evaluated the RADI’s relationship with these existing indices, using a Pearson correlation to measure the linear association between RADI and SVI, and an ordered logistic regression to model the relationship between the index scores and the ordinal RUCC categories. Finally, to test for independent contribution, we used multivariable analysis. Two sets of multiple linear regression models were created: one including both RADI and SVI (using standardized beta coefficients for comparison) and another including both RADI and RUCC (as dummy-coded variables). These models allowed us to determine if RADI remained a significant predictor after controlling for existing measures; that is, if it captured a unique dimension of rural social determinants of health.

## 3.0 Results

### 3.1 Construction Results

The construction of the Rural Adversity and Determinants Index (RADI) was guided by a Principal Component Analysis (PCA) that distilled the 18 individual metrics into a single, coherent dimension of adversity. The first principal component (PC1), which forms the basis of the final RADI score, successfully captured 24.9% of the total shared variance across all variables, representing a substantial underlying construct. An examination of the component loadings for PC1 reveals the specific nature of this adversity. The index is primarily driven by a powerful combination of economic hardship and resource deprivation, with the strongest positive loadings from Median Income (0.414), Poverty (0.401), Food Insecurity (0.401), and No Internet Access (0.357). These indicators form the conceptual core of the RADI, defining rural adversity first and foremost as a condition of profound socioeconomic distress. Following these, a secondary tier of influential variables includes indicators of social vulnerability and infrastructure, such as Percent Non-White (0.269), Unemployment (0.257), No Vehicle Available (0.257), and No Plumbing Facilities (0.232). Notably, several variables had very weak loadings on PC1, indicating they contribute minimally to this primary dimension of adversity, including the proportion of the population aged 65 and over (-0.024), Drinking Water Violations (0.007), and Natural Disaster Risk (0.011). This suggests that while these factors may represent rural challenges, they are not systematically correlated with the core socioeconomic distress that unifies the other variables. While not used in the final index, subsequent components captured more niche dimensions of rurality; for instance, the second component (12.45%) was strongly defined by natural disaster risk in contrast to demographic factors. The selection of PC1 for the RADI is therefore empirically justified, as it represents the broadest and most powerful single dimension of shared socioeconomic disadvantage present in the data.

### 3.2 Predictive Validity of the Rural Adversity and Determinants Index

The initial stage of validation involved assessing the standalone predictive power of the RADI for key health outcomes and comparing its performance to the existing SVI and RUCC measures. A series of single-predictor linear regression models was conducted for five distinct health outcomes. The detailed statistical outputs of these models are presented in Table 1.

**Table 1.**
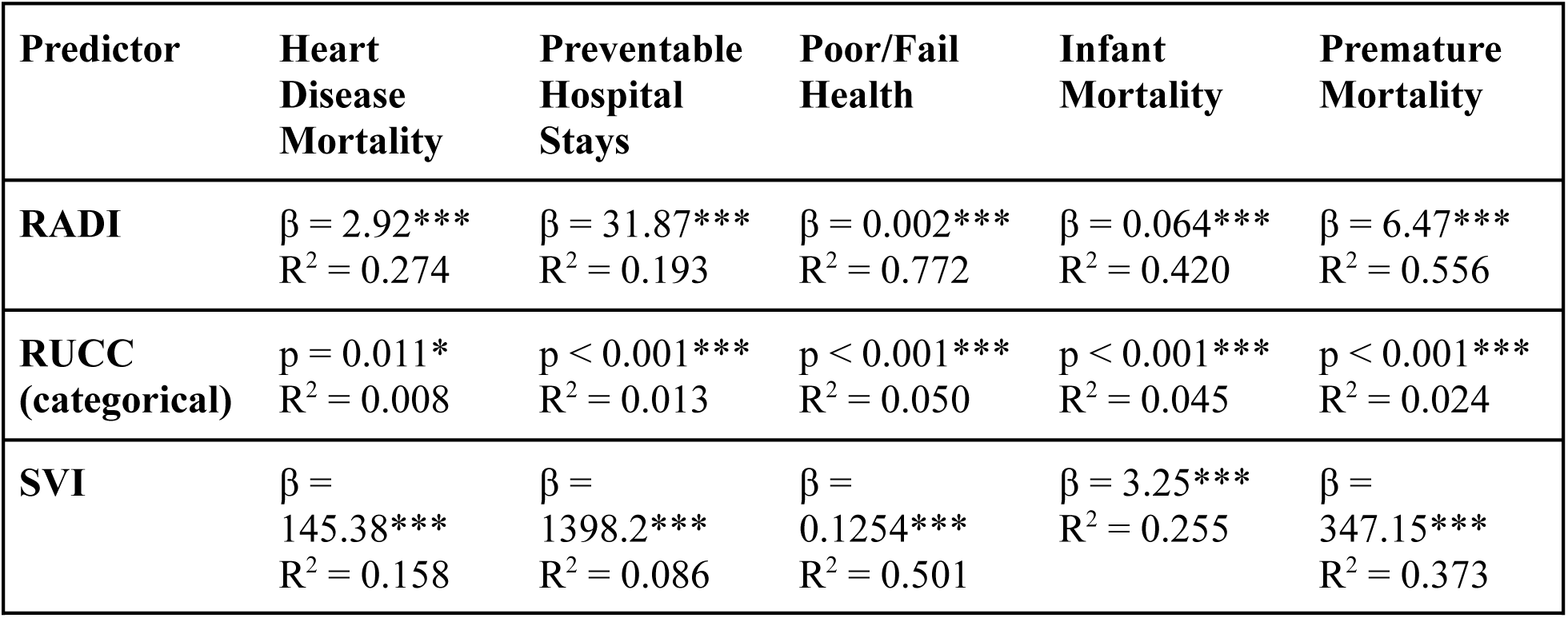
Associations between predictors and health outcomes (Linear Regression results).

The findings from these analyses consistently demonstrated that the RADI score was a robust and statistically significant predictor across all five health outcomes. Furthermore, it generally explained a greater proportion of the statistical variance in these outcomes than either SVI or RUCC. For the outcome of Poor or Fair Health, the RADI explained 77.2% of the variance, a substantial increase over the 50.1% explained by SVI and the 5.0% explained by RUCC. A similar pattern was observed for Premature Mortality, where RADI accounted for 55.6% of the variance, again showing considerably greater explanatory power than SVI (37.3%) and RUCC (2.4%). This trend continued for Infant Mortality (RADI R^2^=0.420 vs. SVI R^2^=0.255), Heart Disease Mortality (RADI R^2^=0.274 vs. SVI R^2^=0.158), and Preventable Hospital Stays (RADI R^2^=0.193 vs. SVI R^2^=0.086). In every case, the RADI outperformed the other indices as a standalone predictor.

The magnitude of these relationships is further clarified by the regression coefficients. For every one-point increase in a county’s RADI score, the rate of premature mortality was associated with an increase of 6.47 years of potential life lost per 100,000 population. The strong positive correlations between the RADI and each of the adverse health outcomes are visualized in the Pearson correlation matrix (Figure 1).

**Figure 1.**
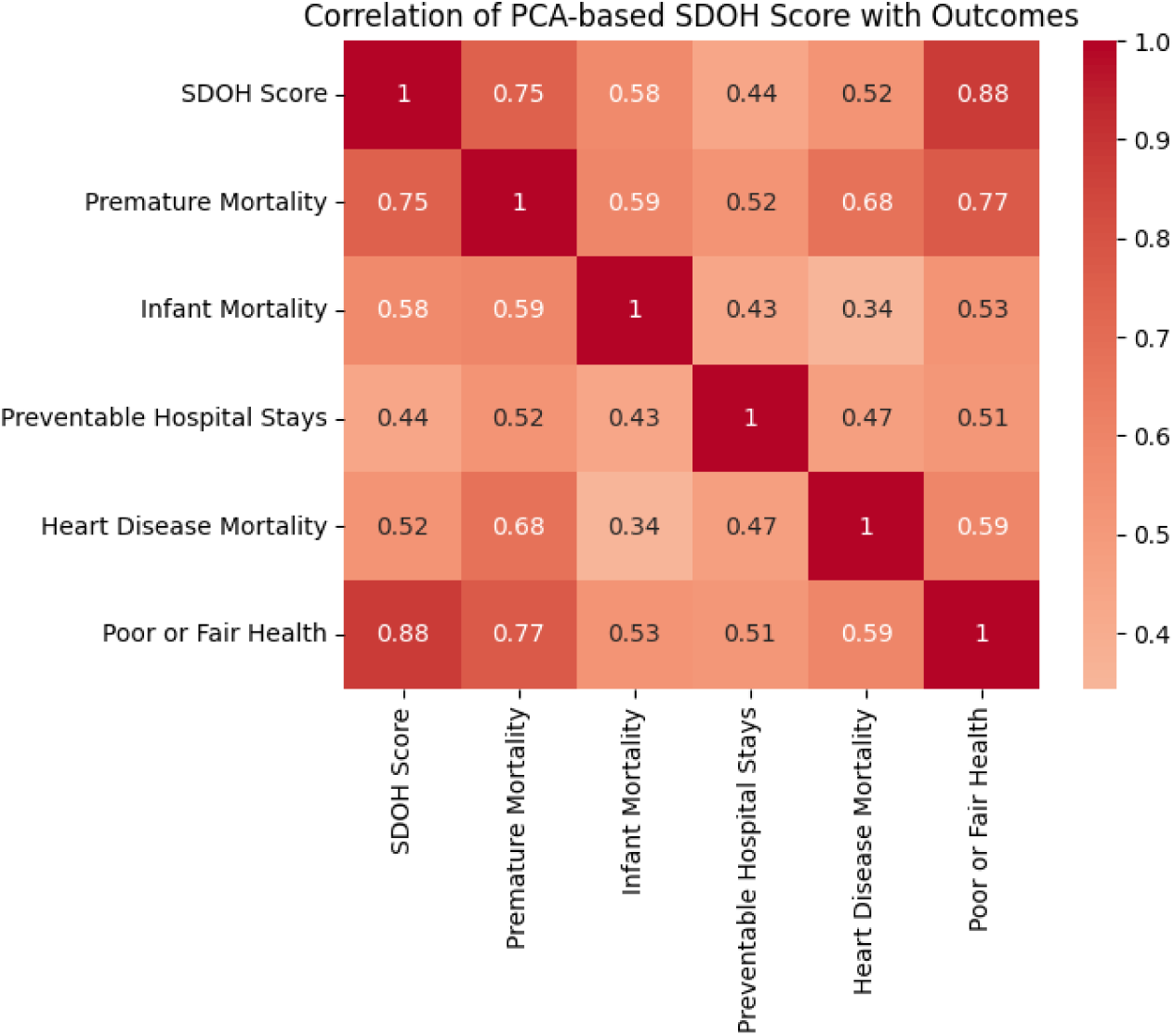
Schematic representation of the Pearson correlation matrix visualizing correlation of PCA-based SDOH score with outcomes.

The geographic distribution of adversity as measured by the RADI is shown in the county-level map. The map reveals distinct spatial clustering, with the highest levels of rural adversity concentrated in specific regions of the country (Figure 2).

**Figure 2.**
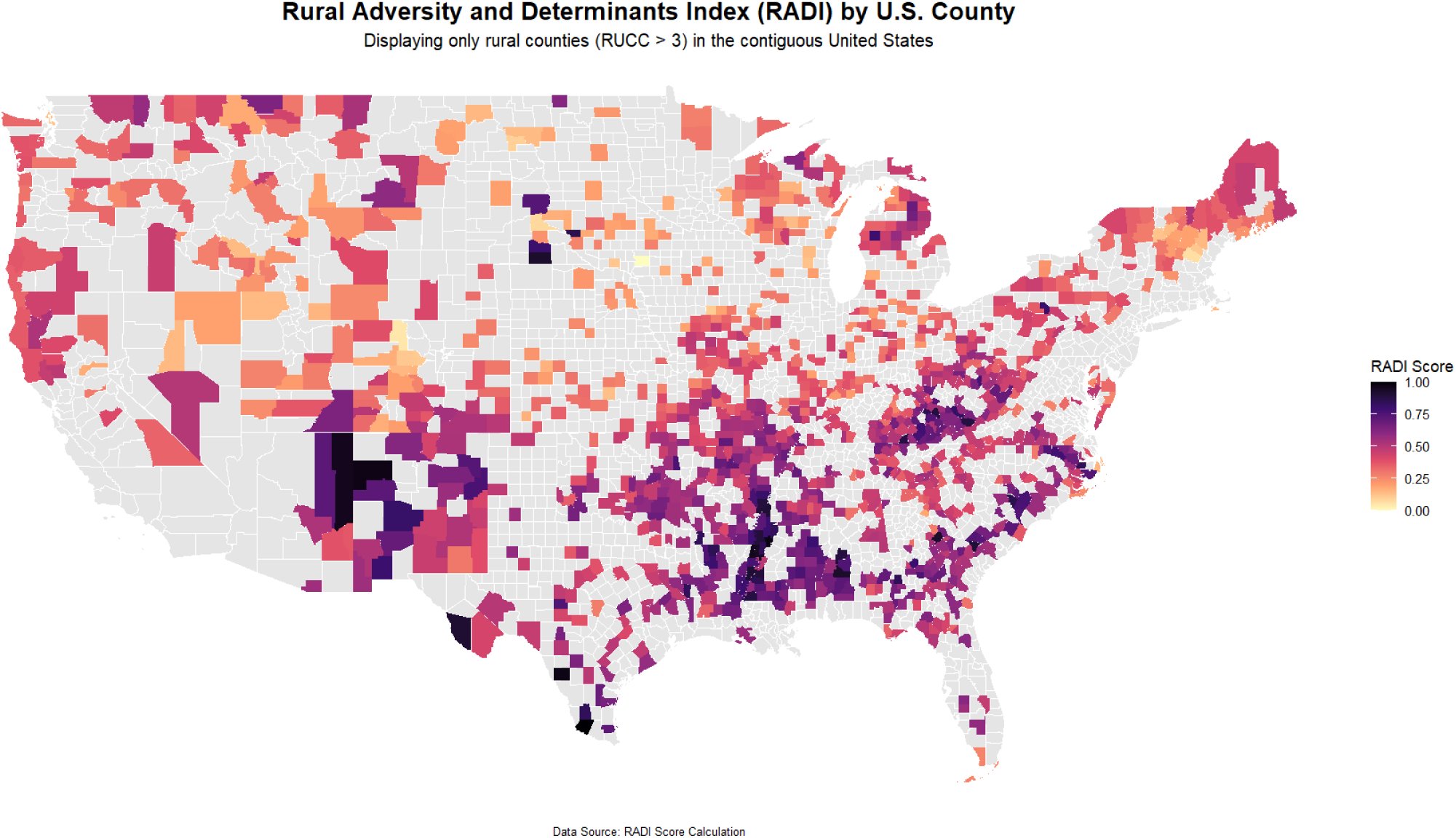
Map depicting the Rural Adversity and Determinants Index (RADI) by U.S. county.

### 3.2 Relationship Between RADI and Existing Indices

To understand how the RADI relates to existing measures, its association with both the SVI and the RUCC system was examined. A Pearson correlation analysis revealed a strong, positive, and statistically significant linear relationship between the RADI and the SVI (r = 0.708). This indicates that the two indices measure related aspects of social vulnerability. However, the fact that the correlation is not perfect suggests that they also capture distinct, non-overlapping dimensions of adversity, supporting the rationale for developing a rural-specific measure.

Further analysis using ordered logistic regression yielded contrasting and insightful findings regarding the indices’ relationship with geographic isolation, with full results available in Table 2.

**Table 2.** Predictors v.s. RUCC (Ordered Logistic Models)

| Predictor | Coefficient ($\beta$) | Significance |
| --- | --- | --- |
| **RADI** | 0.0278 | *** ($p < 0.001$) |
| **SVI** | -0.4279 | *($p = 0.024$) |

Higher RADI scores were found to be significantly associated with higher, more rural and isolated RUCC categories. This result aligns with the theoretical expectation that greater structural adversity would be concentrated in more geographically isolated rural areas. In stark contrast, a counterintuitive relationship was observed for the SVI within this rural-only sample. Higher SVI scores were significantly associated with lower, less isolated RUCC categories. This suggests that the SVI may identify higher vulnerability in larger rural towns rather than in the most remote counties, highlighting a key difference in what RADI and SVI measure within a non-metropolitan context.

### 3.3 Independent Contribution of RADI in Multivariate Models

To test whether the RADI provides unique explanatory power beyond that of existing indices, two sets of multiple linear regression models were estimated.

The first set assessed RADI and SVI as simultaneous predictors of health outcomes. The detailed results, presented in Table 3, show that the RADI remained a highly significant predictor for all five health outcomes even after controlling for the SVI. A comparison of the standardized beta coefficients reveals the relative strength of the RADI. For instance, in the combined model predicting Premature Mortality, the standardized beta for RADI was substantially larger than that for SVI. Critically, for two outcomes, the SVI lost its statistical significance entirely when RADI was included in the model. In the models for Heart Disease Mortality and Preventable Hospital Stays, the SVI coefficient became non-significant, providing strong evidence that the RADI better captures the specific drivers of these outcomes in rural areas.

**Table 3.** Multiple Linear Regression (RADI & SVI)

| Predictor | Heart Disease Mortality | Preventable Hospital Stays | Poor/Fail Health | Infant Mortality | Premature Mortality |
| --- | --- | --- | --- | --- | --- |
| **RADI** | B = 47.24***<br>t = 13.96 | B = 587.62***<br>t = 12.68 | B = 0.04***<br>t = 40.27 | B = 1.00***<br>t = 18.86 | B = 95.29***<br>t = 23.48 |
| **SVI** | B = 5.42 ns<br>t = 1.60 | B = -42.31 ns<br>t = -0.91 | B = 0.01***<br>t = 9.11 | B = 0.16***<br>t = 3.00 | B = 25.34***<br>t = 6.24 |
| **R<sup>2</sup> of Model** | 0.275 | 0.193 | 0.787 | 0.424 | 0.569 |

The second set of models, detailed in Table 4, included RADI alongside the categorical RUCC variable to assess if RADI’s measure of socioeconomic adversity could explain the variance otherwise attributed to geographic classification.

**Table 4.** Multiple Linear Regression (RADI & RUCC)

| Predictor | Heart Disease Mortality | Preventable Hospital Stays | Poor / Fair Health | Infant Mortality | Premature Mortality |
| --- | --- | --- | --- | --- | --- |
| **RADI** | b = 3.03***<br>t = 21.38 | b = 31.94***<br>t = 16.38 | b = 0.002***<br>t = 62.40 | b = 0.06***<br>t = 28.45 | b = 6.60***<br>t = 38.192 |
| **RUCC** | (See Note C) | (See Note A) | (See Note D) | (See Note B) | (See Note A) |
| **R<sup>2</sup> of Model** | 0.281 | 0.193 | 0.776 | 0.429 | 0.558 |
| A) None of the individual RUCC categories show a statistically significant different in outcome compared to the baseline RUCC category (RUCC 4), after accounting for the RADI Score.<br>B) RUCC 6 is the only level with a significant change (b = 0.37, p = 0.004) from baseline compared to RUCC 4 after controlling for RADI score; all other levels had insignificant changes.<br>C) RUCC 8 and 9 are the only levels with a significant change (b = -17.09, p = 0.045, b = -23.61, p = 0.005) from baseline after controlling for the RADI score.<br>D) RUCC 6 is the only level with a significant change (b = 0.007, p = 0.002) from baseline after accounting for the RADI score. |  |  |  |  |  |

RADI remained a highly significant predictor across all five outcome models. The crucial result was that for most outcomes, the RUCC categories themselves became non-significant predictors once the RADI score was controlled for. For example, in the models for Preventable Hospital Stays and Premature Mortality, no individual RUCC category showed a statistically significant difference in the outcome compared to the baseline category after accounting for RADI. This strongly supports the hypothesis that RADI effectively captures the underlying socioeconomic adversity that RUCC only hints at through geography. A few specific exceptions were noted; for instance, RUCC 6 remained a significant predictor for Infant Mortality, and RUCC 8 and 9 remained significant for Heart Disease Mortality. These exceptions suggest that for certain outcomes, specific geographic contexts may harbor unique, unmeasured factors not fully captured even by the comprehensive RADI.

## 4.0 Discussion

### 4.1 Summary of Key Findings

This study successfully developed and validated the Rural Adversity and Determinants Index (RADI), a novel composite measure tailored to assess the multifaceted nature of disadvantage in U.S. rural counties. The results of our validation analyses yield three main findings that advance the understanding of rural health disparities. First, the RADI proved to be a robust and powerful predictor of multiple adverse health outcomes, including premature mortality, poor or fair health, and preventable hospitalizations. In single-predictor models, it consistently explained a greater proportion of the variance in these outcomes than the nationally recognized Social Vulnerability Index (SVI) or a simple geographic classification system (RUCC).

Second, this research reveals that the RADI captures a dimension of rurality that is distinct from both the SVI and from geographic location alone. This distinction is most apparent in the relationship between the indices and escalating rural isolation. While higher RADI scores were associated with more geographically isolated counties, higher SVI scores were, counterintuitively, associated with less isolated rural counties. This suggests the RADI aligns more closely with the lived experience of adversity in the most remote parts of the country.

Third, the social and economic factors synthesized by the RADI appear to be more fundamental drivers of health outcomes than geography itself. This is evidenced by the finding that when the RADI was included in multivariate models, the predictive power of the RUCC classification was often rendered statistically non-significant. This implies that the well-documented “rural penalty” in health is driven less by the fact of a county’s location and more by the specific adverse conditions that are prevalent in that location, which the RADI is designed to measure directly.

### 4.2 Interpretation and Implications of Findings

The strong predictive performance of the RADI can be attributed to several factors inherent in its design. The selection of the 18 input variables was intentionally focused on metrics that reflect the unique challenges of contemporary rural life. Unlike more general indices, the RADI includes specific measures of infrastructure deficits that disproportionately affect rural areas, such as limited internet access – a critical barrier to telehealth and economic opportunity – and drinking water violations, which can signal aging public works systems without a sufficient tax base for upgrades [5, 19]. It also accounts for healthcare access challenges by including ratios of mental health providers, directly addressing the workforce shortages that are a persistent feature of rural healthcare systems [20]. The data-driven Principal Component Analysis methodology further enhances its performance by algorithmically determining the weight of each variable based on the shared patterns in the data, thus avoiding the subjectivity of predetermined weighting schemes [21–22].

The anomalous negative association between the SVI and the RUCC categories within our rural-only sample warrants specific discussion. This finding suggests that the SVI, while effective at a national scale, may not be appropriately specified to differentiate levels of vulnerability within non-metropolitan areas. We hypothesize that this is because the SVI gives significant weight to factors such as racial and ethnic minority status and the proportion of multi-unit housing structures. In many regions, these demographic characteristics are more prevalent in larger, less-isolated rural towns (e.g., RUCC 4 or 5) than in the most remote, sparsely populated, and historically white agricultural counties (e.g., RUCC 8 or 9).

Consequently, the SVI may assign higher vulnerability scores to less-isolated rural areas, while the RADI, with its broader mix of socioeconomic and infrastructural indicators, identifies escalating adversity in the most remote regions, which is likely more reflective of reality.

This leads to a major implication of our work: the conceptual shift from viewing “place” as a simple geographic category to understanding it as a proxy for “condition.” The RADI facilitates this shift by measuring the adverse conditions of a place directly. The focus moves from simply identifying a county as “rural” to quantifying how disadvantaged that rural county is across a spectrum of well-being. This has profound implications for policy, practice, and research. For resource allocation, federal and state agencies could use the RADI to more equitably and effectively target funding for infrastructure projects, healthcare services, and economic development, moving beyond state-level or broad regional grants to county-specific investments based on measured need. Finally, the RADI opens up new avenues for research, allowing investigators to explore the specific mechanisms through which different dimensions of rural adversity influence health outcomes.

### 4.3 Theoretical Integration

The robust performance of the RADI is best understood when situated within several key theoretical frameworks from health geography and medical sociology. These theories provide a deeper understanding of why this place-based index is such a powerful predictor of health outcomes, moving the interpretation beyond statistical association to conceptual significance.

First, our findings critically advance the long-standing ‘composition versus context’ debate [23]. While compositional factors (individual traits) remain vital, RADI’s predictive power demonstrates that the “black box of place effects” – often dismissed as a residual category – holds measurable, contextual drivers of health adversity. Specifically, RADI operationalizes Macintyre et al.’s call to move beyond geographic labels (e.g., RUCC) by quantifying the material infrastructure (e.g., water safety, internet access) and collective social functioning (e.g., community norms, service availability) that define rural adversity [23]. The index’s ability to render RUCC classifications statistically non-significant confirms that the “rural penalty” stems not merely from geography itself, but from the contextual conditions of places – the very “opportunity structures” Macintyre et al. theorized as fundamental to health [23]. This resolves the false dichotomy between composition and context: RADI captures how place characteristics shape both individual outcomes and the collective lived experience of adversity.

The ability of the RADI to predict a wide array of disparate health outcomes -- from infant mortality to preventable hospital stays -- lends strong support to its validity as a measure of fundamental disadvantage. In this, our work aligns with Link and Phelan’s (1995) Fundamental Cause Theory [24–25]. The RADI can be conceptualized as a place-based operationalization of this theory, quantifying the lack of flexible resources (e.g., money, knowledge, power, beneficial social connections) at the community level that are essential for maintaining health. High-RADI counties are not just facing individual health challenges; they are resource-deprived environments where the population is fundamentally constrained in its ability to avoid risks and adopt health-protective behaviors, leading to a pattern of generalized vulnerability across multiple health domains [24–25].

In essence, RADI empirically maps what Gesler (1992) conceptualizes as “therapeutic” and “pathogenic landscapes” [26]. High-RADI regions – with their confluence of economic deprivation, infrastructural decay (e.g., plumbing failures, transportation gaps), and healthcare scarcity – epitomize pathogenic landscapes. These are spaces where material deficits and eroded social foundations (e.g., fragmented community networks, diminished “fields of care”) generate environments hostile to health. Conversely, low-RADI counties align with Gesler’s therapeutic landscapes, where robust infrastructure, accessible services, and cohesive collective functioning create environments conducive to healing. Critically, RADI reveals how these landscapes are not static but socially constructed: the clustering of adversity in Appalachia and the Deep South reflects structural inequities and historical disinvestment that manifest as territorial stigmatization, a key mechanism in Gesler’s framework. This reframes policy from treating individual sickness to transforming pathogenic landscapes by targeting their material and symbolic foundations, such as rebuilding infrastructure and fostering “symbolic pathways” for community resilience [26].

Finally, the structural conditions measured by the RADI likely have profound implications for a community’s social fabric. While not measured directly, it is plausible that the high levels of adversity captured by the RADI erode a community’s ‘collective efficacy’ -- its capacity for social cohesion and mutual trust [27]. The constant struggle with economic instability, resource scarcity, and infrastructure failure can deplete the social capital necessary for residents to organize and advocate for their collective good. Future research could explore this link directly, investigating whether the RADI score predicts measures of social cohesion and civic engagement, thereby uncovering a key social mechanism linking structural adversity to poor health.

### 4.4 Strengths and Limitations

This study has several notable strengths. Its primary strength lies in the novelty of developing a composite index specifically designed for and validated within a large, well-defined sample of U.S. rural counties. The data-driven PCA methodology is another key strength, as it reduces subjectivity in variable weighting and allows the structure of the data itself to define the index. The use of multiple, robust, and publicly available national data sources ensures the index is replicable and can be updated over time. Furthermore, the rigorous validation process, which involved comparison against multiple health outcomes and two established national indices, provides strong evidence for the RADI’s utility and construct validity.

However, the study is not without limitations. First, as an ecological study using county-level data, it is subject to the ecological fallacy; the associations observed at the county level do not necessarily reflect individual-level risk [28]. Second, the cross-sectional nature of the data means that causality cannot be inferred. We can demonstrate a strong association between RADI scores and poor health, but we cannot definitively state that the adversity caused the outcome. Third, there is a potential for data timeliness issues, as some source data, such as the ACS, are based on multi-year survey periods and may lag behind current conditions. Fourth, while the first principal component explained a substantial portion of the variance for a single component in complex social data (approximately 25%), this means that about 75% of the variance is not captured in the final score. This remaining variance likely represents other dimensions of rural life and more localized, idiosyncratic factors that are not systematically shared across all variables. Fifth, rural communities located in non-rural counties (RUCC < 4) are excluded from this index. Finally, the initial choice of the 18 metrics, while theory-driven and comprehensive, is still a choice. The inclusion of other variables could have resulted in a different index structure.

### 4.5 Future Directions

The development and validation of the RADI lays the groundwork for several important avenues of future research. To achieve greater precision for targeting interventions, future work could focus on regional or state indices at smaller units of analyses, such as the census tract, to identify pockets of adversity within counties; such work may help increase the percent variance explained. A longitudinal analysis is also a critical next step. Tracking changes in county RADI scores over time and modeling their impact on health outcomes would allow for a more robust, quasi-causal understanding of the relationship between adversity and health. Moreover, the varying predictive effectiveness with regards to differing health outcomes indicate that purpose-built indices for specific measures (i.e chronic disease, mental health) may prove even more effective.

Furthermore, this quantitative index would be greatly enriched by complementary qualitative research. Conducting interviews and focus groups in counties with particularly high or low RADI scores could provide a deeper understanding of the lived experiences behind the numbers, illuminating the specific community dynamics, policies, and histories that contribute to resilience or vulnerability. From an applied perspective, future research should also include policy impact studies. As agencies potentially begin to use the RADI for resource allocation, it will be essential to track those investments and measure their subsequent impact on health outcomes and equity.

Finally, the index itself could be a subject of ongoing refinement. Future iterations could explore alternative statistical methods, such as factor analysis, or test the inclusion of additional variables related to social capital, civic infrastructure, or climate change vulnerability to potentially increase the explanatory power of the model.

## 5.0 Conclusion

This paper presented the development and rigorous validation of the Rural Adversity and Determinants Index (RADI). The core contribution of this work is the creation of a purpose-built tool designed specifically to fill a critical gap in rural health research and policy. The RADI is not merely another index of social vulnerability; it is a nuanced instrument constructed from the ground up to measure the unique combination of social, economic, and infrastructural challenges that define adversity across the diverse landscape of rural America. Our findings demonstrate that the RADI effectively captures these distinct dimensions of disadvantage and, as a result, serves as a powerful predictor of health disparities, often outperforming broader, national-level indices within a rural context.

By providing a more accurate and granular measure of which rural places are struggling the most and why, the RADI offers a new lens for understanding and addressing long-standing inequities. The ultimate potential of this tool lies in its application. By enabling more precise targeting of resources, informing the design of tailored public health interventions, and opening new avenues for research, the RADI can contribute directly to the creation of more equitable health outcomes and improved well-being for the millions of Americans who call rural communities home.

## Statements and Declarations

The authors disclose no conflicts of interests or other declarations.

## Data Availability

All data produced in the present study are available upon reasonable request to the authors

